# Preceding anti-spike IgG levels predicted risk and severity of COVID-19 during the Omicron-dominant wave in Santa Fe city, Argentina

**DOI:** 10.1101/2022.05.18.22275138

**Authors:** Ayelen T. Eberhardt, Melina Simoncini, Carlos Piña, Germán Galoppo, Virginia Parachú-Marco, Andrea Racca, Sofía Arce, Evangelina Viotto, Florencia Facelli, Florencia Valli, Cecilia Botto, Leonardo Scarpa, Celina Junges, Cintia Palavecino, Camila Beccaria, Diego Sklar, Graciela Mingo, Alicia Genolet, Mónica Muñoz de Toro, Hugo Aimar, Verónica Marignac, Juan Carlos Bossio, Gustavo Armando, Hugo Fernández, Pablo M. Beldomenico

## Abstract

The SARS-CoV-2 Omicron variant has increased infectivity and immune escape compared with previous variants, and caused massive COVID-19 waves globally. Despite a vast majority (∼90%) of the population of Santa Fe city, Argentina, had been vaccinated and/or had been infected by SARS-CoV-2 by the arrival of Omicron, the epidemic wave that followed was by far the largest one experienced in the city. A serosurvey conducted prior to the arrival of Omicron allowed to assess the acquired humoural defences preceding the wave and to evaluate associations with infection risk and severity during the wave. A very large proportion of the 1455 sampled individuals had immunological memory against COVID-19 at the arrival of Omicron (almost 90%), and about half (48.9%) had high anti-Spike IgG levels (>200 UI/ml). The antibody titres were strongly associated with the number of vaccine shots and the vaccine platform received, and also depended markedly on prior COVID-19 diagnosis and the days elapsed since last antigen exposure (vaccine shot or natural infection). Following-up 514 participants provided real-world data linking antibody levels with protection against COVID-19 during a period of high risk of exposure to an immune-escaping highly transmissible variant. Pre-wave antibody titres were strongly associated with COVID-19 incidence and severity of symptoms during the wave. Also, receiving a vaccine shot during the follow-up period reduced the COVID-19 risk drastically (15-fold). These results highlight the importance of maintaining high defences through vaccination anticipating or during a period of high risk of exposure to immune-escaping variants.

## Introduction

As of June 2022, the COVID-19 pandemic continues to occur despite the acquired defences developed in a large proportion of people due to vaccination against SARS-CoV-2 and/or natural infection. Several viral variants have evolved, prevailing the ones that achieved enhanced transmissibility and immune escape compared to prior variants (Tian et al. 2022). Until November 2021, some strains had become prominent and had caused new outbreaks worldwide. These were considered variants of concern, and were named Alpha, Beta, Gamma, and Delta. A new variant, B.1.1.529, was first detected in samples collected on 11^th^ November 2021 in Botswana and on 14^th^ November 2021 in South Africa (Tian et al. 2022). On 26^th^ November, the WHO defined it as the fifth variant of concern, naming it Omicron. So far, Omicron is the variant with the largest number of mutations, many of which provide increased infectivity and immune escape compared with previous variants (Hu et al. 2022; Duong et al. 2022). This resulted in massive waves of COVID-19 emerging worldwide soon after the new variant appeared (Daria & Islam, 2022).

The dynamics of COVID-19 have been heterogeneous since the beginning of the pandemic (Beldomenico 2020). While countries like United Kingdom and Germany have gone through several epidemic waves, others like Thailand and Vietnam had their first wave only after over a year had passed since SARS-CoV-2 began to circulate in those countries. In Argentina, by early December 2021 there had been two waves, the first one by mid-2020, related to the arrival and spread of the virus, and the second one in 2021 associated with the seasonality of respiratory viruses. Omicron was confirmed in Argentina on 5^th^ December 2021, and some days later the country suffered the largest COVID-19 epidemic wave so far, with a peak infection rate several times higher than the peaks observed in the two previous waves.

In Santa Fe, a city of around 430,000 inhabitants, COVID-19 dynamics reflected what was observed elsewhere in the rest of the country (Figure 1). By mid-December 2021, 12.9% of the citizens had been diagnosed with COVID-19, 90.6% had received a first dose of an anti-SARS-CoV-2 vaccine, 79.2% a second dose, and 10.3% a third one (data provided by the Ministry of Health of Santa Fe province). The wave that followed the arrival of Omicron began around 18^th^ December 2022 in Santa Fe city, and the number of daily cases started to decline by mid-January 2022, returning to levels as low as before the wave by the end of February (Figure 1).

**Figure 1.**
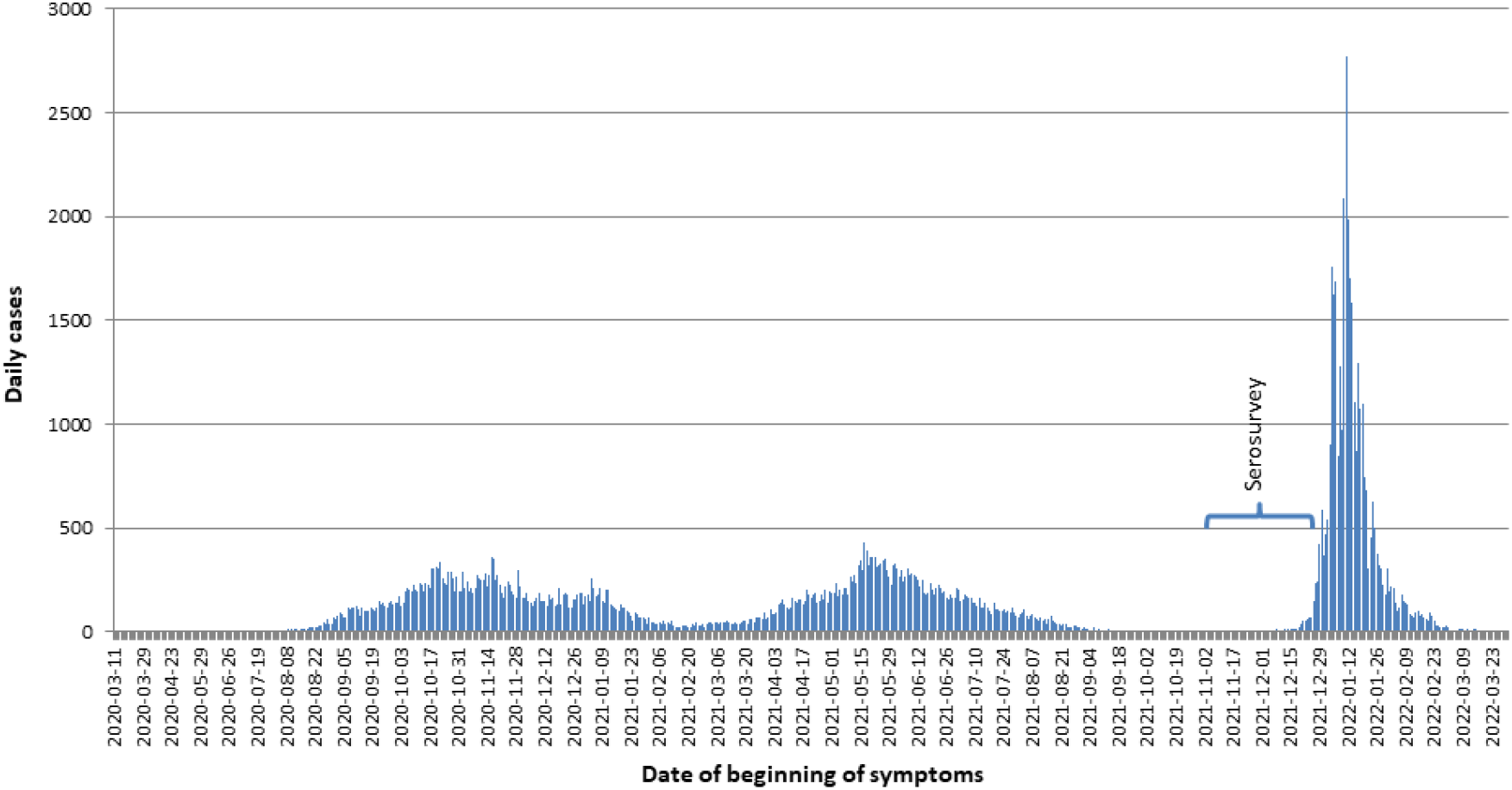
Temporal distribution of the confirmed cases of COVID-19 in Santa Fe city (official records of the Ministry of Health of Santa Fe province).

During the last two months of 2021, we conducted a survey collecting relevant information on COVID-19 and measuring anti-spike IgG antibodies in people from randomly selected households of Santa Fe city and from citizens that volunteered to participate in the study. This provided the opportunity of characterising the acquired humoural defences of the population of Santa Fe city immediately prior to the arrival of Omicron. In March 2022, after the wave was over, a subset of the study participants were asked to complete a second questionnaire indicating if they were diagnosed with COVID-19 after 15th December 2021, if they got additional vaccine shots, and other relevant information. The data collected allowed us to pursue three goals:

1. to describe the acquired humoural immunity of the population immediately prior to the arrival of Omicron>
2. to assess which factors were associated with such immune status (i.e. previous infection, different vaccination schemes, time from last exposure, etc.), and>
3. to evaluate if those humoural defences predicted the risk and severity of COVID-19 during the wave.>

## Materials and methods

### Source of the data

A random sample of 1000 households including all neighbourhoods of Santa Fe city was provided by the Instituto Provincial de Estadísticas y Censos. In September and October 2021, those households were visited, and the occupants were invited to participate in a COVID-19 study that involved answering a questionnaire and providing a blood sample to measure IgG antibodies against SARS-CoV-2. A second visit was scheduled from 1^st^ November 2021 to 23^rd^ December 2021, to fill the questionnaires and take a blood sample. In addition, during that period volunteers were invited to participate by announces in the local media. We collected data from 414 people from randomly selected households and 1041 volunteers.

The first questionnaire included queries on sex, age, having been diagnosed with COVID-19 (with dates and diagnosis details), COVID-19 severity and duration, vaccine shots received (with type and dates), close contacts with COVID-19 cases, co-morbidities, among other information.

Those that were sampled after 15^th^ November 2021, were asked to complete a second questionnaire in March 2022. This allowed us to follow the participants from whom there was an antibody measurement within a month prior to the Omicron-dominant wave. The second questionnaire inquired for the period that went from the date of the blood sample collection to 28^th^ February 2022, and included information on close contact with cases during that period, COVID-19 diagnosis (with dates and diagnosis details), vaccine shots (with type and dates), and disease severity and duration. When there were doubts about the responses given in the questionnaires, the participants were contacted again to ask for clarification. We obtained 514 valid responses to this second questionnaire.

Using the information obtained from the questionnaires, we established a COVID-19 case when the participant indicated that she/he was given positive by the government after an official PCR test, or given positive by the government due to having symptoms while cohabitating with a case, or had a PCR positive test by a private laboratory, or had a positive rapid antigen test while having symptoms or after having had a close contact with a case. People that declared they suspected having had COVID-19 but were not tested (25 in the first questionnaire and 4 in the second questionnaire) were removed from the analysis.

All procedures were carried out under the approval of the Ethics and Biosafety Committee of the Scientific and Technological Centre of Santa Fe of the Argentine Council for Research and Technology (CCT Santa Fe CONICET). All participants signed an informed consent.

### Quantification of IgG

Levels of anti-SARS-CoV-2 spike protein IgG were quantified by COVID AR IgG immunoassay developed by Instituto Leloir in Argentina (Ojeda et al. 2021), following the manufacturer’s instructions. This IgG immunoassay kit consists of a solid phase ELISA that utilizes the trimer of native protein S and a receptor binding domain as antigens, obtained by recombinant DNA techniques produced in human cells.

Briefly, 40 µl of fingertip capillary blood samples were diluted 1:6 in the diluent provided in the SEROKIT developed by Instituto Leloir, and kept refrigerated. At the laboratory, samples were re-diluted 1:3, and 200 µl of each 1:18 final dilution were transferred to 96-well plates and incubated at 37°C for one hour. IgG specific for spike protein was captured on the plate, and subsequently the wells were thoroughly washed 6 times to remove unbound material. Anti-Human IgG, HRP-linked antibody was then used to recognize the bound IgG. A mix of HRP substrate and TMB (1:1) was added to develop color. The magnitude of optical density at 450 nm is proportional to the quantity of IgG specific for spike protein. To estimate antibody levels, sample optical densities were converted to concentrations expressed in UI/ml by using a lineal model built with the optical densities (response variables) obtained in each plate from two sets of known dilutions of the positive control at 50, 100, 200 and 400 UI/ml. These dilutions were the independent variable, included as a polynomial term (with lineal and quadratic terms) to address possible non-linearity of the dilution-OD relationship. The R^2^ of that model was checked to confirm that the value was >0.85, otherwise all samples were analysed again in a new plate.

### Statistical analysis

All statistical analyses were done using the Software R version 4.2.0 (The R Foundation for Statistical Computing). The analyses were conducted in three steps, to pursue three complementary goals, as follows.

The first step aimed to characterise the acquired humoural defences in Santa Fe immediately prior to the arrival of Omicron. This part consisted of descriptive statistics of the IgG levels, overall and by age group.

The goal of the second step was to investigate the determinants of the IgG levels measured. For this, the antibody levels were the response variable, which were transformed by calculating the square root to approach normality. Two sets of lineal models were run, one containing the polynomial term ‘number of doses + (number of doses)^2^’ as variable of interest (to take into account possible non-linearity of the titre-dose relationship), and the second set including only the 4 vaccination schemes most frequently observed, to compare antibody levels among them. In both models, the independent variables ‘COVID-19’ (prior diagnosis of COVID-19) and ‘days from last exposure’ (vaccine shot or detected infection; whatever happened last), were included. The vaccination schemes used for the second model were: two Astra-Zeneca vaccines (viral vector vaccine; N= 411), two Sinopharm (inactivated vaccine; N= 334), two Sputnik V vaccines (viral vector vaccine; N= 260), the combination of Spunik V and Moderna (viral vector + mRNA vaccines; N= 155), and two Pfizer/BioNTech (mRNA vaccine; N= 25).

The third step used information from the second questionnaire to conduct a longitudinal analysis that enabled assessment of how the vaccines and antibody levels influenced the incidence of COVID-19 during the Omicron-dominant wave in Santa Fe. In addition, we looked at associations between antibody levels and COVID-19 symptoms severity and duration among those that were infected during the Omicron-dominant wave. For this third step, the period in which participants were followed to assess new detected infections by SARS-CoV-2 was from December 18^th^ to February 28^th^ (72 days).

In order to establish an association between vaccination status and the incidence of COVID-19 during the Omicron-dominant wave, we built a Generalized Lineal Models (GLM) with a binomial response (COVID-19 positive or not). The model used number of vaccine doses as the independent variable of interest, included as a polynomial term.

To assess associations between antibody levels and the incidence of COVID-19 during the Omicron-dominant wave, we used a subset of data (N= 484) that excluded participants that got a vaccine shot between 7 days prior of the blood sample collection and 15th December 2021 (N= 30), as for those cases the antibody levels recorded did not reflect the humoural immunity on the arrival of Omicron. We built a GLM with a binary response that evaluated associations between antibody levels, while adjusting by a number of relevant factors, detailed below. The independent variable of interest (IgG levels) was also included in a separate model as a dichotomous factor, setting those with antibody levels >400 UI/ml (very high levels; N= 158) as 1, and the rest as 0 (N= 326).

Finally, in the subset of samples that was diagnosed with COVID-19 during the wave (N= 184) the associations between antibody levels and COVID-19 severity and duration were assessed with ordinal regression models, where the responses were 3-level ordinal variables, as follows. Disease severity was measured by asking in the second questionnaire whether they had no or very mild symptoms (e.g. light sore throat, nasal congestion; level 1), mild symptoms (e.g. one or two days of fever and/or light malaise, not requiring bed rest; level 2), or moderate symptoms (e.g. bed rest was required; level 3). The participants were also asked if hospitalization was required, as a 4^th^ level, but none chose this option. As for the duration of COVID-19 symptoms (excluding loss of smell), the three levels were: one day or less (level 1), two to five days (level 2), and more than 5 days (level 3).

For all models used in step 3, potential confounding phenomena was controlled for by including in the models relevant independent variables, as follows. Age (in years, and assessed separately as a single term or polynomial) was included in all models. Also in all models, receiving a new vaccine during the wave period was included as a two-level independent variable, as those that got a booster shot within the follow-up period had changes in both the antibody levels and the vaccination scheme. The cases in which the vaccine shot was received late in the wave (after 15^th^ February 2022; N= 7) were not used in the models that included the new shot variable. The number of known close contacts with COVID-19 cases was used as a proxy of exposure, and used for the GLMs assessing associations with COVID-19 incidence. Close contact was defined as being within 3 m distance or indoors for over 15 minutes with someone who was diagnosed with COVID-19, and the contact happened within the period that went from two days prior the onset of symptoms and seven days after the onset of symptoms. The contacts were set at 5 levels, 0, 1, 2, 3, and 4 or more close contacts with cases. Prior COVID-19 was included in models that assessed associations between vaccination status and COVID-19 incidence during the wave. Finally, the presence of co-morbidities (i.e. high blood pressure, diabetes, obesity, heart disease, chronic pulmonary disease, cancer) was included in the models assessing the influence of antibody levels on COVID-19 severity and duration. All these variables used for adjustment purposes were dropped from the models if they were not important for the model’s goodness of fit, as indicated by AIC comparisons.

## Results

### Description of the sample

We obtained answers to the first questionnaire and blood samples from 1455 people. Of those, 57.3% were female and 43.7% were male. The mean age was 41 years old, being the minimum 5 months old and the maximum 95 years old.

Almost three quarters (74.7%) of the participants had not been diagnosed with COVID-19 at the time of answering the first questionnaire, but 2.4% of those (N= 25) suspected having been infected. One quarter (24.8%) was diagnosed with COVID-19 once, and 0.4% (N= 6) twice.

Regarding the vaccination regime, 6.9% of the participants were not vaccinated at the time of sampling, 5% had one dose, 83.6% had two doses, 4.5% had three doses and 0.07% had four doses. The vaccination scheme most frequently applied in the sample was two Astra-Zeneca vaccines, followed by two Sinopharm, two Sputnik V vaccines, and the combination of Spunik V and Moderna. At the time of the sampling, Pfizer/BioNTech vaccines were being used for youngsters aged 13 to 18 years old, having 25 participants of our study two doses, and 19 one dose.

### Characterisation of the acquired humoural defences prior to the Omicron-dominant wave

Anti-spike IgG were detected in 88.7% (1290/1455) of the samples. Among those that received at least one dose of an anti-COVID-19 vaccine, 7.4% (100/1354) did not have detectable IgG. Among the non-vaccinated (N=99), 63.6% (63/99) did not have detectable antibodies. Of the unvaccinated that had antibodies, 72% (26/36) had not been diagnosed with COVID-19 nor suspected having been infected.

Almost half (48.9%) of the participants had antibody levels considered to be high (>200 UI/ml), and more than one third (35.4%) had very high levels (>400 UI/ml). The overall mean antibody level was 290 UI/ml, but it varied by age group (Table 1). Among the age group considered to be of high risk (>60 years old), the vast majority was vaccinated (98.1%), but 17% was vulnerable because they had no detectable IgG (6%) or had low antibody levels (11% with <40 UI/ml). However, most aged 60 and above had very high antibody levels (65% with >400 UI/ml). The high level of antibodies observed in those aged 13-20 is attributable to the good performance of the vaccine received by that age group and the shorter time elapsed from the last shot.

**Table 1.** Central tendency (mean and median) of antibody levels and proportion of vaccine coverage (at least one shot) by age group, in samples taken from Santa Fe citizens in November and December 2021.

| Age range | Sample size | Mean; Median (UI/ml) | % without antibodies | % vaccinated |
| --- | --- | --- | --- | --- |
| 0-12 years old | 84 | 175; 12 | 43.4% | 47.0% |
| 13-20 years old | 87 | 474; 499 | 9.0% | 85.4% |
| 21-40 years old | 535 | 217; 82 | 12.2% | 95.8% |
| 41-60 years old | 438 | 307; 223 | 9.5% | 96.0% |
| > 60 years old | 311 | 378; 408 | 5.9% | 98.1% |

### Antibody levels according to vaccine doses and schemes

Those participants that were not vaccinated had a mean IgG titer of 62.5 UI/ml, while the mean level was 287.9 UI/ml, 293.5 UI/ml, and 567.8 UI/ml for those that received one, two or three doses, respectively. A third dose increased the antibody levels significantly, and there was strong positive association with prior COVID-19 and a strong negative correlation with days that elapsed from last exposure (vaccine or infection) (Figure 2, Table 2).

**Table 2:** Lineal model assessing the association between antibody (IgG) levels and number of anti-COVID-19 vaccine doses received, adjusting by prior COVID-19 diagnosis and days from last exposure (vaccine or known infection). Significant terms are printed in bold.

| Term | Coefficients | Standard error | P-value |
| --- | --- | --- | --- |
| **Intercept** | **15.6487** | **1.6665** | **<0.0001** |
| **Vaccine doses** | **-1.7496** | **1.5259** | **0.2518** |
| **Vaccine doses<sup>2</sup>** | **1.2979** | **0.4477** | **0.0038** |
| **COVID-19** | **5.0559** | **0.5401** | **<0.0001** |
| **Days from exposure** | **-0.0388** | **0.0036** | **<0.0001** |

Antibody levels: Titre of anti-SARS-CoV-2 spike protein IgG.
Vaccine doses: number of vaccine shots received prior to the blood sample.
COVID-19: prior diagnosis of COVID-19.
Days from exposure: days elapsed from the last vaccine shot or detected infection .

**Figure 2.**
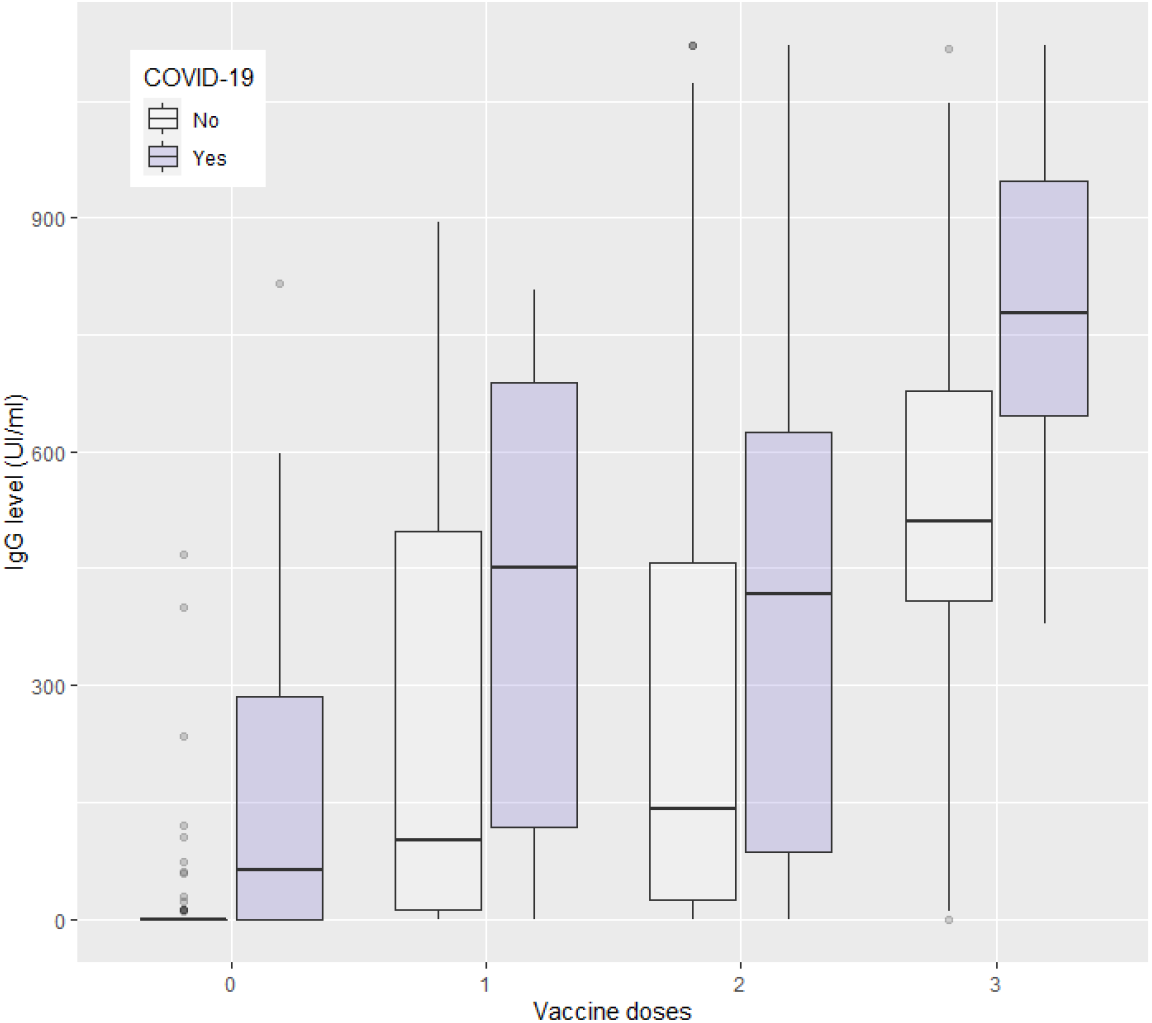
Levels of antibodies (IgG) against SARS-CoV-2 by number of vaccine doses and prior COVID-19 diagnosis.

When comparing the four vaccination schemes most frequently applied while adjusting by prior COVID-19 infection and days elapsed from last exposure, we observed very significant differences in antibody levels (Table 3; Figure 3). The scheme with inactivated vaccines (Sinopharm) showed significantly lower antibody levels than all other schemes, both schemes of viral vectors (Astra-Zeneca and Sputnik V) performed similarly, and the schemes combining vector and mRNA (Sputnik V + Moderna) and two mRNA (Pfizer/BioNTech) showed the highest levels, not statistically different between them (Table 3).

**Table 3:**
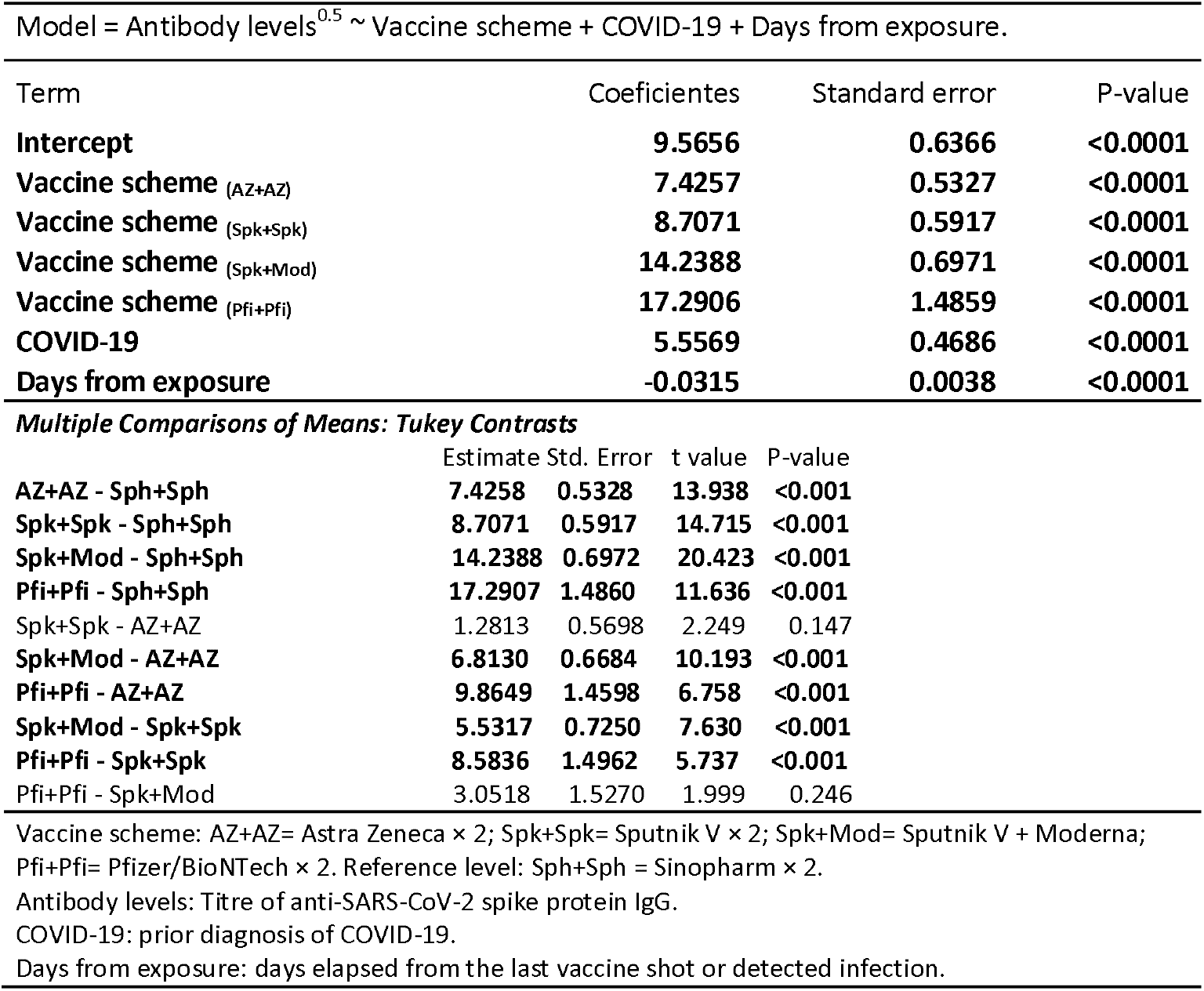
Lineal model assessing the association between antibody (IgG) levels and different anti-COVID-19 schemes, adjusting by prior COVID-19 diagnosis and days from last exposure (vaccine or known infection). PostHoc Tukey tests indicated the significant differences between vaccination schemes. Significant terms are printed in bold.

**Figure 3.**
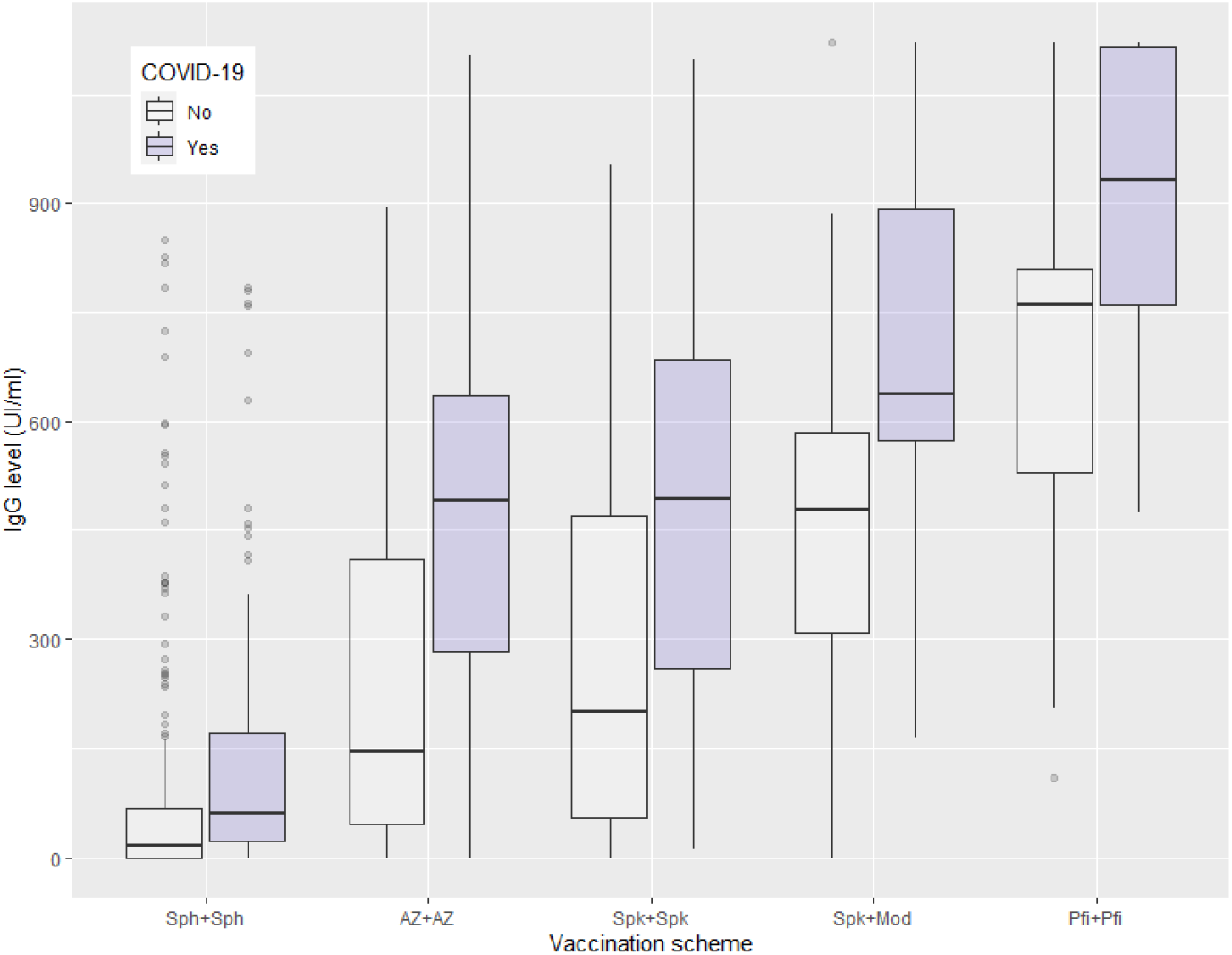
Levels of antibodies (IgG) against SARS-CoV-2 by vaccine scheme and prior COVID-19 diagnosis. Sph= Sinopharm; AZ= Astra Zeneca; Spk= Sputnik V; Mod= Moderna; Pfi= Pfizer/BioNTech

### Vaccination status and COVID-19 incidence during the Omicron-dominant wave

Of the 514 participants followed up during the wave, 184 (35.8%) were diagnosed with COVID-19 between 18^th^ December 2022 and 28^th^ February 2022. The incidence in those that had had COVID-19 previously was also high (reinfection rate: 39/121 = 32.2%). The number of vaccine doses received prior to the arrival of Omicron did not appear to have an effect on the COVID-19 infection risk during the wave (Table 4). However, a deeper analysis taking into account age, prior COVID-19, receiving a vaccine dose during the wave, and close contact with cases, showed that the number of doses was significantly associated with infection risk in a non-linear way, as observed for the association with antibody levels (Table 2), indicating that a significant reduction in infection risk was observed only for those with 3 or more vaccine shots (Table 5). Contact with cases and vaccination during the wave were strong predictors of COVID-19 risk during the wave, and prior COVID-19 was also significantly associated (Table 5).

**Table 4.** COVID-19 attack rate during the Omicron-dominant wave in Santa Fe city, by number of vaccine doses received prior to the onset of the wave.

| Number of vaccine doses: | 0 | 1 | 2 | 3 | 4 |
| --- | --- | --- | --- | --- | --- |
| N= | 41 | 9 | 405 | 58 | 1 |
| Diagnosed with COVID-19= | 15 | 6 | 146 | 17 | 0 |
| Attack rate= | 36.6% | 66.7% | 36.0% | 29.3% | - |

**Table 5.** Logistic regression assessing the association between COVID-19 diagnosis (yes/no) during the Omicron-dominant wave and number of vaccine doses received before the wave, adjusting by age, prior COVID-19, vaccine shot during the wave and number close contacts with cases. Significant terms are printed in bold.

| Model = COVID-19 (yes/no) ~ Age + Vaccine doses + Vaccine doses<sup>2</sup> + Prior COVID-19 + Contact + New vaccine shot |  |  |  |
| --- | --- | --- | --- |
| Term | Coefficients | Standard error | P-value |
| Intercept | 0.4628 | 0.0713 | <0.0001 |
| Age | -0.0009 | 0.0011 | 0.4330 |
| **Vaccine doses** | **0.3299** | **0.0704** | **<0.0001** |
| **Vaccine doses<sup>2</sup>** | **-0.1166** | **0.0229** | **<0.0001** |
| **Prior COVID-19** | **-0.1050** | **0.0417** | **0.0122** |
| **Contact** | **0.0896** | **0.0212** | **<0.0001** |
| **New vaccine shot** | **-0.5161** | **0.0386** | **<0.0001** |
| Age: in years. |  |  |  |
| Vaccine doses: number of vaccine doses received before the wave (0 - 4). |  |  |  |
| Prior COVID-19: diagnosis of COVID-19 before the Omicron-dominant wave. |  |  |  |
| Contact: number close contacts with cases during the Omicron wave. |  |  |  |
| New vaccine shot: vaccine shot during the Omicron wave. |  |  |  |

### Antibody levels and COVID-19 incidence during the Omicron-dominant wave

There was a strong negative association between antibody levels preceding the Omicron-dominant wave and COVID-19 incidence during the wave (Figure 4, Table 6 and Table S1). For every 100 UI/ml increase in IgG levels, the risk of infection decreases 12%, adjusting by recent vaccine shot and contact with cases (Table 6). Participants with antibody levels >400 UI/ml at the onset of the wave had 67% less chances of being diagnosed with COVID-19 during the wave (Table S1 in supplementary material). In addition, receiving a vaccine shot after the onset of the wave and the number of close contacts with cases were strong predictors of COVID-19 risk in all models. A vaccine shot during the wave reduced 15-fold the probability of COVID-19 (Table 6). Every close contact with a case increased 71% the odds of being diagnosed with COVID-19 during the wave (Table 6).

**Table 6.** Logistic regression assessing the association between COVID-19 diagnosis (yes/no) during the Omicron-dominant wave and antibody levels, adjusting by vaccine shot during the wave and number close contacts with cases. Significant terms are printed in bold.

| Model = COVID-19 (yes/no) ~ Antibody levels + New vaccine shot + Contact |  |  |  |
| --- | --- | --- | --- |
| Term | Coefficients<br>(log-odds) | Standard error | P-value |
| Intercept | 0.3857 | 0.2050 | 0.0599 |
| **Antibody levels** | **-0.0011** | **0.0004** | **0.0062** |
| **New vaccine shot** | **-2.7105** | **0.2661** | **<0.0001** |
| **Contact** | **0.5349** | **0.1419** | **0.0001** |
| Antibody levels: Titre of anti-SARS-CoV-2 spike protein IgG. |  |  |  |
| New vaccine shot: vaccine shot during the Omicron wave. |  |  |  |
| Contact: number of close contacts with cases during the Omicron wave. |  |  |  |

**Figure 4.**
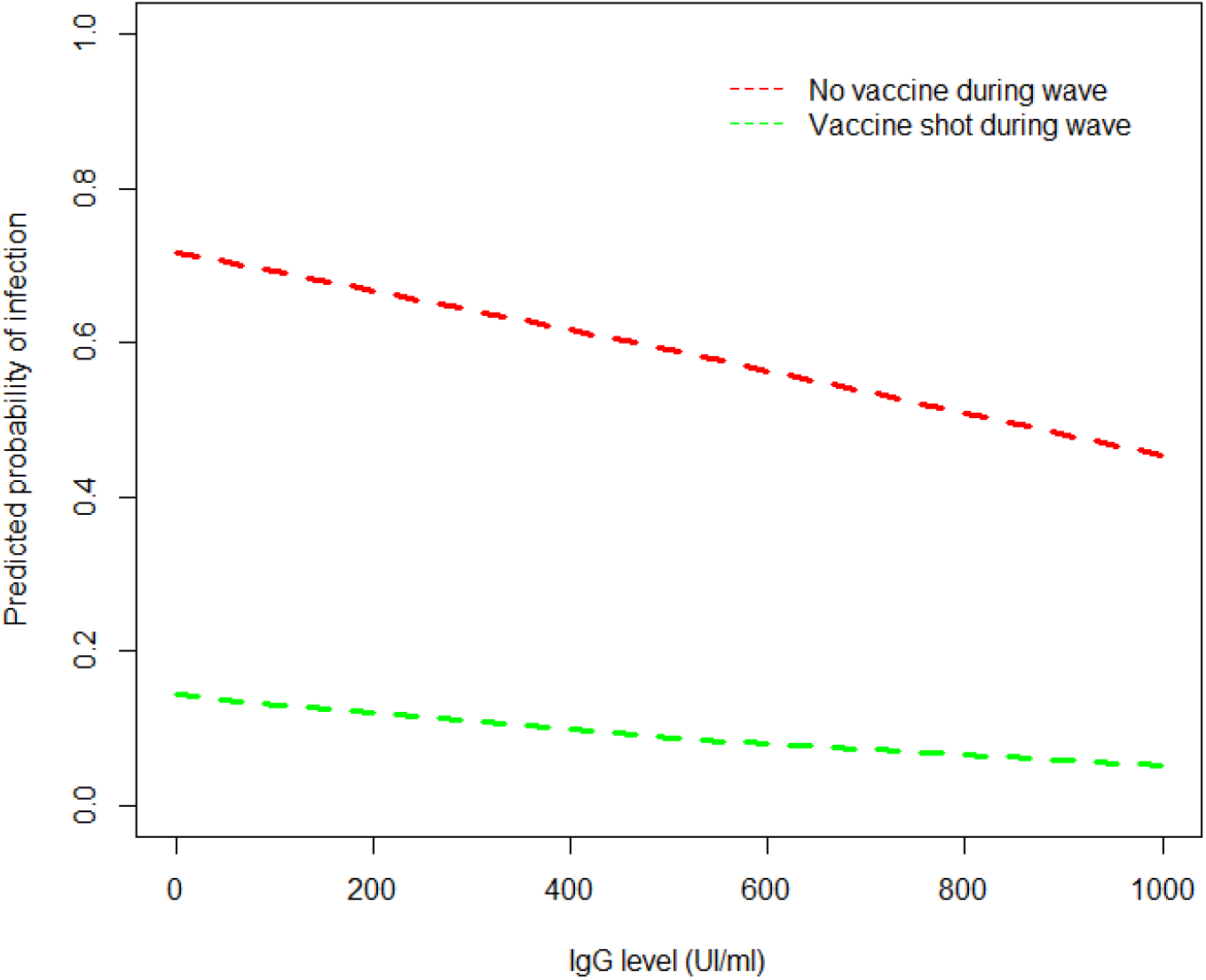
Predicted probability of COVID-19 during the Omicron-dominant wave depending on the levels of antibodies (IgG) against SARS-CoV-2 and the administration of a vaccine dose during the wave. For the simulation contact with cases was set at 1.

### Antibody levels and COVID-19 severity and duration

The ordinal regression model showed that antibody levels were strongly associated with the severity of the symptoms (Table 7, Figure 5). For every 100 UI/ml increase in the IgG level, the odds of being more likely to have higher disease severity (mild or moderate symptoms versus none or very mild symptoms) decreases 34.8%, holding constant new vaccine shot, age and presence of co-morbidities. The model looking at the association between antibody levels and duration of the symptoms showed a negative trend, but not statistically significant, although borderline (p=0.05; Table S2 in supplementary material).

**Table 7.** Ordinal regression model assessing the association between severity of COVID-19 symptoms and antibody levels, adjusting by vaccine shot during the wave, age and co-morbidities. Significant terms are printed in bold.

| Model = Severity ~ Antibody levels + New vaccine shot + Age + Co-morbidity. |  |  |  |
| --- | --- | --- | --- |
| Term | Coefficients | Standard error | P-value |
| **Antibody levels** | **-0.0019** | **0.0005** | **<0.0001** |
| **New vaccine shot** | **-1.0189** | **0.4580** | **0.0261** |
| Age | -0.0089 | 0.0112 | 0.4235 |
| Co-morbidity | -0.7914 | 0.4744 | 0.0953 |

Severity: severity of COVID-19 symptoms; 3-level ordinal variable: very mild symptoms, mild symptoms or moderate symptoms. Antibody levels: Titre of anti-SARS-CoV-2 spike protein IgG. New vaccine shot: vaccine shot during the Omicron wave. Age: in years. Co-morbidity: conditions like high blood pressure, diabetes, obesity, heart disease, chronic pulmonary disease, cancer.

**Figure 5.**
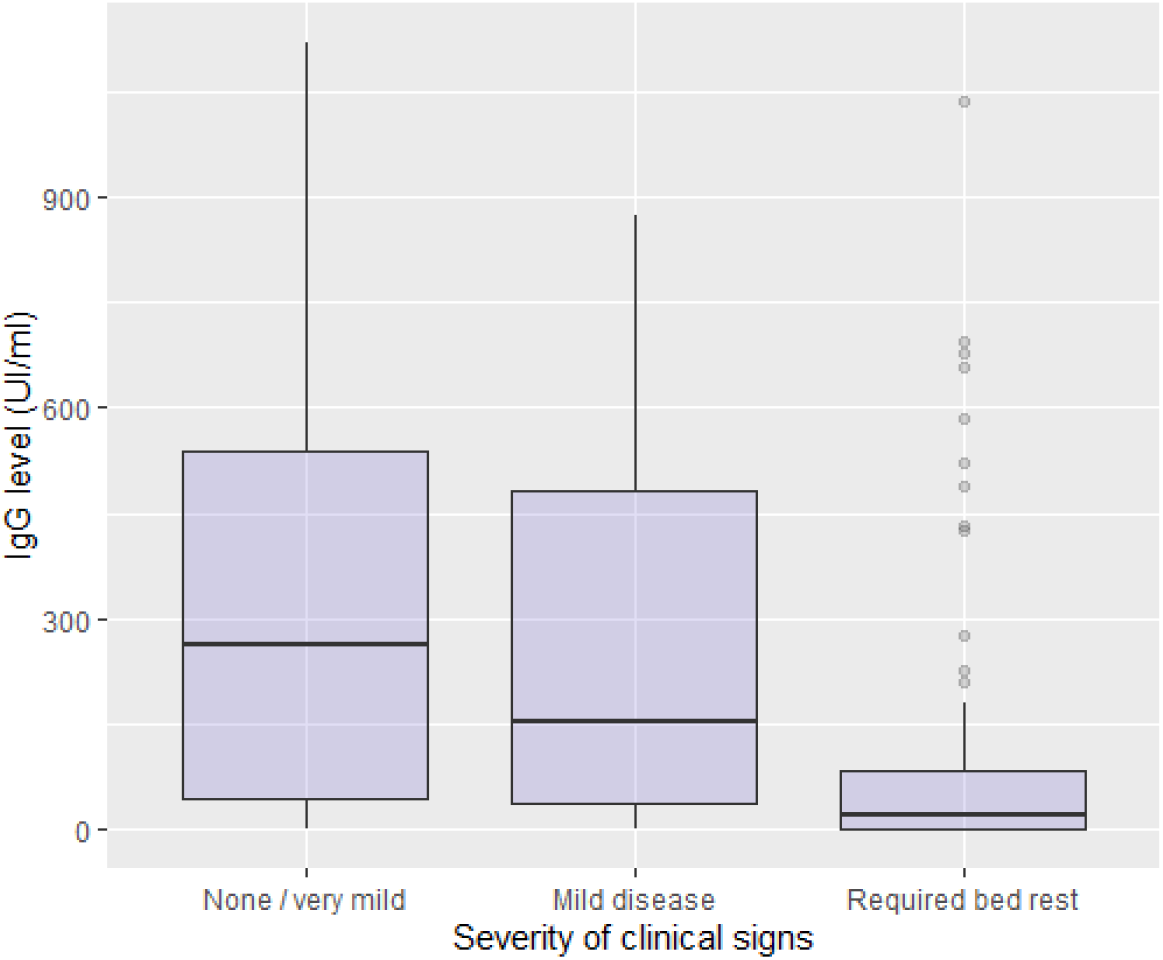
Levels of antibodies (IgG) against SARS-CoV-2 at the onset of the Omicron-dominant wave by the severity of the symptoms when they became infected during the wave. None or very mild symptoms, N= 59; mild disease, N= 69; disease that required bed rest, N= 59.

## Discussion

The arrival of the variant Omicron was associated with the largest wave of COVID-19 cases in Santa Fe city (Figure 1), despite immediately prior to the wave a vast majority of the citizens (>90%) had been vaccinated and/or had been infected by SARS-CoV-2. Here we characterised the acquired humoural defences in Santa Fe prior to the arrival of Omicron, estimating that a high proportion of the population had immunological memory against COVID-19 (i.e. almost 90% of our sample had detectable antibodies). Moreover, about half of the participants had high antibody levels (>200 UI/ml). Although the number of cases during the Omicron-dominant wave was much higher than in the previous two waves, the mortality due to COVID-19 was considerably lower. Before the Omicron-dominant wave, there had been 856 deaths over 55969 cases (case-fatality= 1.5%) in Santa Fe, and during the wave there were 54 deaths over 36166 cases (case-fatality= 0.15%) (official records of the Ministry of Health of Santa Fe province). This 10-fold lower impact could be attributable to the high level of defences here described, which is supported by the individual-level data presented, where the severity of COVID-19 was lower the higher the preceding antibody levels. However, because there is evidence that suggests that Omicron may be less pathogenic than previous variants (Ulloa et al. 2022; Wolter et al. 2022) it is difficult to infer how much of the reduced severity is due to immunological experience and how much attributable to virus evolution making new variants less pathogenic (Bhattacharyya et al. 2022).

Antibodies provide protection either through direct obstruction of infection or through their ability to leverage the immune system to eliminate pathogens. The neutralising antibody titres generated in vaccine clinical trials are assumed to be correlated with protective effect and the durability of the protection (He et al. 2021). Measuring antibody-mediated protection to coronaviruses requires characterisation of immune responses prior to a known exposure or period of risk. Such data is only available from few human challenge experiments, in which volunteers were exposed to experimental infections with human coronaviruses (Huang et al. 2020). Some of those studies showed evidence that pre-exposure titres correlated negatively with infection risk and severity (Bradburne et al. 1967; Callow 1985). More recent relevant evidence comes from treatments with convalescent plasma. The efficacy of convalescent plasma transfusion as a treatment for COVID-19 was found to depend on the antibody levels of the plasma. Plasma with higher anti–SARS-CoV-2 IgG antibody levels was associated with a lower risk of death (Joyner et al. 2021). In this longitudinal study, we provide individual-level real-world data linking antibody levels and protection against COVID-19 during a period of high risk of exposure to an immune-escaping highly transmissible variant.

We found that the level of antibodies of the participants immediately prior to the arrival of Omicron were strongly associated with the number of vaccine shots and the vaccine platform received, and also depended markedly on prior COVID-19 diagnosis and the days elapsed since last exposure (vaccine shot or infection). Those that received a third vaccine dose had much higher antibody levels than participants that got 2 or less shots at the time of sampling, which resulted from the booster effect already documented (Kanokudon et al. 2022). It is noteworthy that in our study this large difference between the titres of 2 and 3 shots was maintained when adjusting by days from the last shot.

Another factor that explained the variability in antibody levels in our sample was the vaccination scheme. The vaccine schemes used in Argentina showed different performance in terms of immunogenicity. Two inactivated vaccines (Sinopharm) conferred the lowest antibody levels, and schemes that used mRNA platforms (Sputnik + Moderna or Pfizer/BioNTech × 2) the highest titres, whereas both vector vaccines (Astra Zeneca × 2 or Sputnik V × 2) performed between the other two schemes. This is in agreement with what was reported previously (Banki et al. 2022; Kanokudon et al. 2022; Kudlay & Svistunov 2022).

Despite the strong association between vaccination status and antibody levels, an association of the former with COVID-19 incidence during the wave was only apparent when adjusting by age, prior COVID-19, number of close contacts and vaccination during the wave. When all those factors were held equal, only those with 3 doses were found to have lower chances of being diagnosed with COVID-19 during the wave. A recent study showed that the neutralization potency against Omicron was undetectable in sera from most vaccinees, except for individuals recently receiving a RNAm vaccine booster (3^rd^ dose) (García-Beltran et al. 2022). The apparent lack of effect observed in those with 1 or 2 vaccine doses might result from the large variability in immunogenicity of the vaccines used and the strong correlation observed with days elapsed from the last vaccine shot (as those with 3 shots had been vaccinated more recently). It was documented that anti-spike IgG wane quickly (Bayart et al. 2021; Levin et al. 2021), and here we confirmed this in a real-world study and showed consequences of waning defences on disease risk and severity.

Prior studies have shown increased antibody evasion and greater breakthrough infection risk of Omicron, compared with previous variants (Hoffmann et al. 2021; Mannar et al. 2022; Hu et al. 2022). However, although reduced, the binding of IgG antibodies to the Omicron Spike antigen is maintained, and recent data suggests that extraneutralising antibodies contribute to disease control (Bartsch et al. 2022). This partial immune escape implicates that higher defence levels would be required to reduce the risk and severity of COVID-19 caused by the Omicron variant. Here we present evidence of this in real circumstances. Anti-spike IgG levels and variables that cause antibodies to rise (i.e. prior COVID-19 and a recent boost shot; Table 5) were strong drivers of COVID-19 risk and severity. Our results strongly suggest that to reduce the impact of highly transmissible and immune-escaping variants like Omicron, the acquired defences should be kept high. Therefore, booster vaccine shots anticipating or during a period of high exposure risk are highly recommended.

The results hereby presented offer an explanation to the epidemiological pattern observed in Santa Fe city during the Omicron-dominant wave. The arrival of the Omicron variant caused the largest COVID-19 epidemic experienced in Santa Fe city since the beginning of the pandemic, but the case-fatality observed was 10-fold lower than that of previous waves. The increased number of cases may be have been caused by the immune escape and high transmissibility of Omicron while the high immune defences existing in the population at the time of its arrival most likely contributed the low impact observed. Disease risk and severity was lowest in individuals with high antibody levels, which highlight the importance of maintaining high defences through vaccination in the presence of immune-escaping variants.

## Data Availability

All data produced in the present study are available upon reasonable request to the authors

## Acknowledgements

This work was funded by the Agencia Santafesina de Ciencia, Tecnología e Innovación (Grant # DEMES-2020-0008). The Instituto Provincial de Estadísticas y Censos of Santa Fe province provided the random sample of households. Laboratorio Lemos, Instituto Leloir, Dr. Andrea Gamarnik and Dr. Marcelo Yanovsky donated part of the assays used in this work. Special thanks to all participants for accepting taking part in this study. To visit the selected households and collect the samples, we got assistance from Georgina Brusco, Marina Visconti, Triana Rodriguez, Shirley Musio, Julieta Maldonado, María Laura Arce, María Belén Marinaro, Lucía Slaboch, Lucía Jalit, Camila Maldonado, Florencia Bergogne Cis, Maira Gutierrez, Matías Palmero, Karen Mendoza, Antonella Menegazzo, Rocía Gareis, Emiliano Grandoli, Ivana Ondarcuhu, Camila Zlauvinen, Damián Doña, María Rocío Bustos, Eduardo Masat and Juliana Torancio.

