## Supplementary tables (1 and 2) for "Preceding anti-spike IgG levels predicted risk and severity of COVID-19 during the Omicron-dominant wave in Santa Fe city, Argentina"

Supplementary material

**Table S1**. Logistic regression assessing the association between COVID-19 diagnosis (yes/no) during the Omicron-dominant wave and antibody levels as a dichotomous variable (>400 UI/ml; ≤400 UI/ml [reference]), adjusting by vaccine shot during the wave and number close contacts with cases. Significant terms are printed in bold.

| Model = COVID-19 (yes/no) ~ High antibody levels + New vaccine shot + Contact. | | | |
| --- | --- | --- | --- |
| Term | Coefficients  (*log-odds*) | Standard error | P-value |
| Intercept | 0.2503 | 0.1897 | 0.1870 |
| **High antibody levels** | **-0.5115** | **0.2444** | **0.0364** |
| **New vaccine shot** | **-2.6941** | **0.2659** | **<0.0001** |
| **Contact** | **0.5358** | **0.1408** | **0.0001** |
| High antibody levels: dichotomous variable of the titre of anti- SARS-CoV-2 spike protein IgG: 0= ≤400 UI/ml) [reference]; 1= >400 UI/ml.  New vaccine shot: vaccine shot during the Omicron wave  Contact: number of close contacts with cases during the Omicron wave. | | | |

**Table S2**. Ordinal regression model assessing the association between duration of COVID-19 symptoms and antibody levels, adjusting by vaccine shot during the wave, age and co-morbidities. Significant terms are printed in bold.

| Model = Duration ~ Antibody levels + New vaccine shot + Age + Co-morbidity. | | | |
| --- | --- | --- | --- |
| Term | Coefficients | Standard error | P-value |
| Antibody levels | -0.0012 | 0.0006 | 0.0508 |
| New vaccine shot | -0.7247 | 0.5334 | 0.1743 |
| **Age** | **0.0300** | **0.0143** | **0.0366** |
| Co-morbidity | -1.0919 | 0.6163 | 0.0764 |
| Duration: duration of COVID-19 symptoms (excluding loss of smell). Three levels: one day or less (level 1), two to five days (level 2), and more than 5 days (level 3).  Antibody levels: Titre of anti-SARS-CoV-2 spike protein IgG.  New vaccine shot: vaccine shot during the Omicron wave.  Age: in years.  Co-morbidity: conditions like high blood pressure, diabetes, obesity, heart disease, chronic pulmonary disease, cancer. | | | |
